# Personalizing the Pressure Reactivity Index for Quantifying Cerebral Autoregulation in Neurocritical Care

**DOI:** 10.1101/2023.05.08.23289682

**Authors:** Jennifer K. Briggs, J.N. Stroh, Brandon Foreman, Soojin Park, the TRACK-TBI Study Investigators, Tellen D. Bennett, David J. Albers

## Abstract

**Objective:** The Pressure Reactivity Index (PRx) is a common metric for assessing cerebral autoregulation in neurocritical care. This study aimed to improve the clinical utility of PRx by enhancing its robustness through the development of a personalized PRx algorithm (pPRx) and the identification of ideal hyperparameters.

**Methods:** Algorithmic errors were quantified using simulated and multimodal monitoring data from traumatic brain injury patients from the TrackTBI dataset. Using linear regression between errors and physiological quantities, heart rate was identified as a potential cause of PRx error. The pPRx method was developed by reparameterizing PRx averaging to heartbeats. Ideal hyperparameters for the standard PRx algorithm were identified that minimized algorithmic errors.

**Results:** The PRx algorithm was highly sensitive to hyperparameters and patient variability. Errors were closely related to patient heart rates. By parameterizing PRx to heartbeats, the pPRx methodology significantly reduced sensitivity to both patient variability and hyperparameter selection, while also decreasing noise. In the standard PRx algorithm, averaging windows of 10 seconds and correlation windows of 40 samples resulted in the lowest overall error.

**Conclusion:** Personalized PRx enhances the robustness and accuracy of cerebral autoregulation estimation by addressing patient- and hyperparameter-sensitivity. This improvement is crucial for reliable clinical decision-making in neurocritical care.

**Significance:** Robust estimation of cerebral autoregulation functionality would be beneficial for identifying precision medicine targets and improving outcomes for neurocritical care patients. We systematically increased the robustness of PRx, a common current metric of cerebral autoregulation functionality, to make it more consistent across patient populations.

## I. Introduction

Neurological injuries are a prevalent cause of long-term disability and death. In the United States, there are approximately 64,000 traumatic brain injury (TBI)-related deaths[1] and 160,000 stroke-related deaths[2] annually. Poor patient outcomes often result from secondary insults relating to inadequate perfusion. Therefore, optimizing perfusion is a primary objective in neurocritical care.

Cerebral perfusion, typically represented by its proxy, cerebral blood flow (CBF), depends on cerebral vascular resistance (CVR) and cerebral perfusion pressure (CPP). CPP is the pressure gradient formed between mean intracranial pressure (mICP) and mean arterial blood pressure (mABP) (**eq. 1**). Current clinical guidelines recommend targeting mICP below 22 mmHg to mitigate the risk of secondary injuries[3]. However, a precision medicine approach that considers the state of the patient’s vasculature may improve patient outcomes.

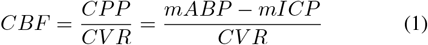

Cerebral autoregulation (CA) is an intrinsic mechanism of the cerebral vasculature that maintains CBF over large pressure gradients by altering CVR[4], [5]. CA is often impaired in neurological injury[6]. When CA is impaired, clinicians must carefully manage perfusion pressure to prevent ischemia. A precise and reliable measure of CA functionality would aid decision-making in neurocritical care by assessing a patient’s cerebral hemodynamic response to pressure changes and guiding personalized treatment through personalized perfusion targets based on CA functionality.

The most common metric to assess CA function in neurocritical care is the pressure reactivity index (PRx)[5], [7], [8]. PRx is calculated by first applying a non-overlapping moving average to ICP and ABP time series over a predefined window and then computing the Pearson correlation coefficient between a specified number of these averaged ABP and ICP samples[9] (**Fig. 1**). Because they are chosen *a priori*, the size of the averaging and correlation window serve as hyperparameters. Values for these hyperparameters vary in literature (see **Table I**), and the impact of hyperparameter choice is not well understood.

**Fig. 1.**
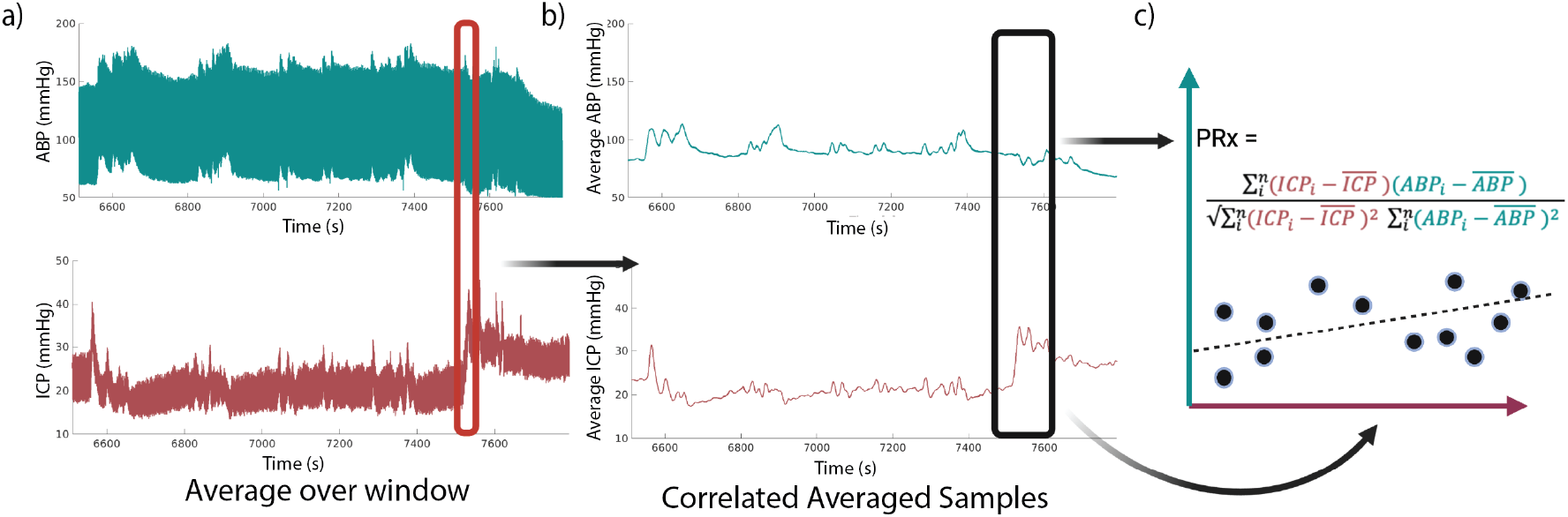
Standard Algorithm for Calculating PRx. a) ABP (blue) and ICP (red) are averaged over a non-overlapping averaging window. b,c) A given number of these averaged samples are collected, and the Pearson correlation coefficient is calculated on a 4/5 overlap sliding window.

**TABLE I.** Common hyperparameters used to calculate PRx. ^*^Howells recommends using 15-50 second averages but does not include a correlation window. We chose correlations that would result in five minute samples consistent with [15], [16].

| Averaging Window (seconds) | Number of Samples for Correlation |
| --- | --- |
| 10 | 40[10] |
| 10 | 30 (most common[11])[12]–[19] |
| 5 | 40[7], [9], [20]–[22] |
| 15 | 30[23]* |
| 6 | 40[8], [24], [25] |

This study aims to enhance the robustness of the PRx algorithm by identifying ideal hyperparameters and developing a personalized parameterization. Hyperparameters could have a substantial impact on PRx calculation robustness because computed quantities (e.g. averages and correlations) are dependent on the number of samples in the calculation. Heart rate directly affects the number of heartbeats, and consequently, the number of samples within the window used to average ICP and ABP, which in turn induces variability in the PRx calculation. Therefore, we test the hypothesis that heart rate affects errors in PRx and that a PRx algorithm personalized to heart rate will provide a more robust and stable estimate of CA functionality.

## II. Methods

### Data

#### Patient Data

Continuous arterial blood pressure and intracranial pressure data recorded at 125 Hz were collected from patients enrolled in the Transforming Research and Clinical Knowledge in Traumatic Brain Injury (TRACK-TBI) study[26], ClinicalTrials.gov ID: NCT02119182. TRACK-TBI is a prospective, multicenter study of patients with traumatic brain injury. Written informed consent was obtained, and the study was approved by the institutional review boards of enrollment sites.

Two datasets termed “development” and “validation” datasets, were extracted (**Table II**). The development dataset included 21 four-hour windows of data (which is the time recommended to calculate a target cerebral perfusion pressure (CPPopt)[21]) from 11 patients from a single center to ensure standardized physiological measurements and clinical practice. The validation dataset included 10 four-hour windows from 8 additional patients from two separate centers to ensure that our results were not dependent on the hospital.

**TABLE II.**
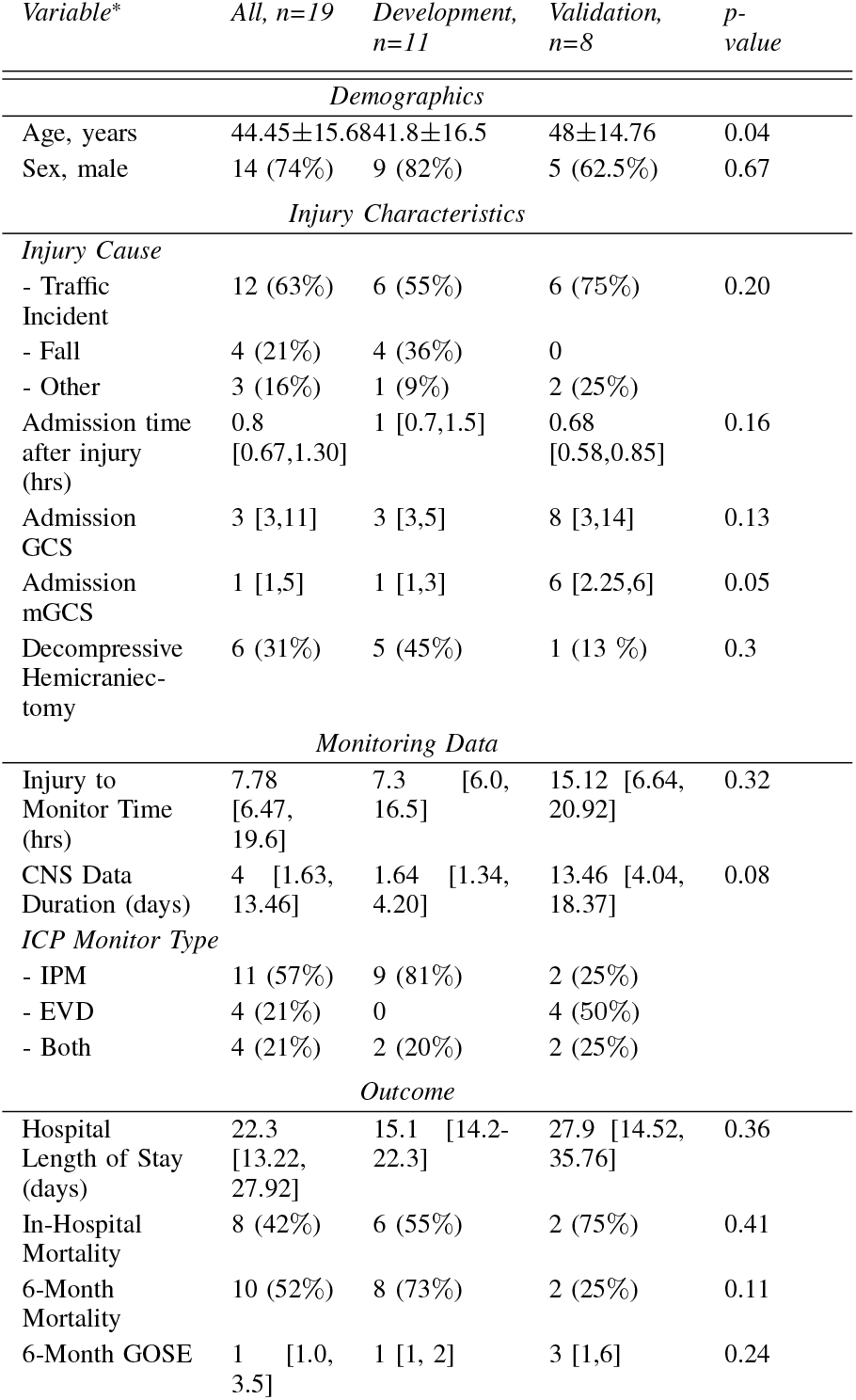
Patient Characteristics and Monitoring Data. The injury was closed for all patients. GCS stands for Glasgow Coma Scale, and mGCS stands for motor Glasgow Coma Scale. IPM stands for Intraparenchymal Monitor. EVD stands for External Ventricular Drain. GOSE stands for Glasgow Outcome Scale-Extended. P-value was calculated using Chi-squared (categorical data), Kruskal–Wallis (non-normal distributions), or t-test (normal distributions) to ensure statistical similarity between groups. All data reported as mean+/-standard deviation, median [interquartile range], or proportion (**%**) as appropriate

To mitigate the impact of missing data on our analyses, datasets with more than 10% of missing or erroneous data were not included. Missing data accounted for 1.7% ± 1.7% of the development data set and 0.5% ± 0.7% of the validation data set. Physiologically implausible values (*>*400 or *<*0 for ABP and *>*100 or *<*0 for ICP) were set to null. Segments of null data shorter than one second were linearly interpolated to fill in a portion of the pressure waveform. This interpolation did not notably impact the PRx estimate. When missing data segments were longer than one second, we omitted the averaging and correlation calculations that would have included these missing data segments.

#### Simulated Data

Continuous evaluation of global CA function in humans is not currently possible. Therefore, validating which hyperparameter would give the true PRx estimate and calculating PRx estimation error is infeasible for patient data. To test PRx errors, a simulated dataset was created based on physiologically relevant frequencies.

ABP reflects physiological components such as heart rate and respiration, as well as other variables such as patient movement. To simulate ABP, four sine waves with frequencies corresponding to these components were combined (**Fig. 2, Table III**). Morphological features of the arterial pulse wave form were not modeled because they are shorter than the smallest averaging window in our analysis and will always be smoothed out. Two hundred twenty four-hour timeseries were created (200 for the development data set and 20 for validation), each with a heart rate randomly selected between 40-140 beats per minute.

**Fig. 2.**
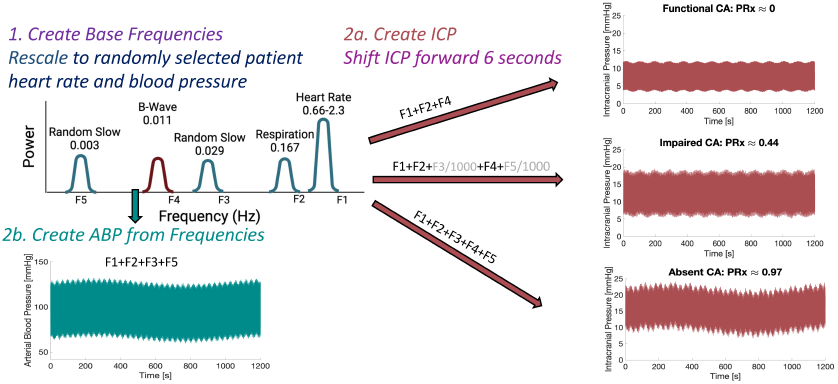
Simulated Data Set Development. Step 1 shows the power spectrum of the six base frequencies (Table III) used to create ICP and ABP. Blue shows a representative ABP timeseries. Red indicates three ICP timeseries created from combinations of the base frequencies. Three CA phenotypes were created: Functional CA, Impaired CA, and Absent CA. The equations used to create each ICP waveform are given above the red arrows

**TABLE III.** Physiologically relevant frequencies used in simulated data.

| Signal Name | Interpretation | Rate or Period |  |
| --- | --- | --- | --- |
| F1 | Heart Rate | 40-140 beats/minute | Scaled in frequency and amplitude to match randomly selected patient |
| F2 | Respiration | 10 breaths/minute |  |
| F3 | Random | 30 second period | Random external changes to the patient[31], [32] |
| F4 | Frequencies only present in ICP | 90 second period | Found only in ICP[27] |
| F5 | Random | 16-minute period | Slow random external changes to the patient |

ICP consists of both independent components and influences from ABP, depending on CA functionality[27]. For each ABP timeseries, three ICP timeseries were created to reflect three CA phenotypes: Functional, Impaired, and Absent. CA phenotypes were based on experimental evidence that when CA is functional, ICP primarily reflects high-frequency components, and as CA functionality declines, lower-frequency components from ABP progressively influence ICP[7], [15], [28], [29]. The equations defining each CA phenotype are provided in **Fig. 2**. ICP timeseries were shifted by 6.8 seconds to simulate the lag behind ABP[30].

To account for variability in the magnitude of ICP and ABP values across patients, joint gamma distributions were fit to characterize the ICP and ABP measurements of 104 patients in the TrackTBI cohort. The ABP and ICP timeseries for each simulated patient were then rescaled to align with the distributions of a randomly selected patient.

PRx is meant to capture the correlation between ABP and ICP after removing fast frequencies that CA does not operate on[31]. Therefore, ‘true’ PRx was defined as the correlation coefficient between simulated ABP and ICP with heart rate and respiration frequencies removed.

### Analysis

Definitions and interpretations of the metrics used to quantify error and sensitivity of the PRx calculation are given in **Table IV**.

**TABLE IV.** Interpretation of Metrics Used to Evaluate PRx Performance.

| Evaluation Metric | Interpretation |
| --- | --- |
| PRx Estimation Error | Difference between PRx estimate from a single hyperparameter compared to true PRx |
| Standard Deviation (SD) or Range of Error | How much patient variability influences error |
| Fine-grained Sensitivity: $\Delta PRx / \Delta \text{window}$ | PRx sensitivity to hyperparameters or missing data (see discussion) |
| Course-grained Sensitivity: Range of PRx values | How much PRx can change across the five common hyperparameters |
| Noise | Noise in PRx estimate |

#### PRx Calculation

To quantify sensitivity to hyperparameters, PRx was calculated using all combinations of averaging windows between 1 and 30 seconds and correlation windows between 2 and 65 averaged samples. This broad range was chosen to capture all hyperparameters present in the literature.

According to standard methods, averaging windows did not overlap, and correlation windows overlapped by 4/5[15]. Unless otherwise noted, the reported PRx value is the median PRx across the four-hour dataset. Here, the notation Avg: x seconds, Corr: y samples is used to indicate the hyperparameter pair with an averaging window of x seconds and correlation window of y samples.

#### PRx Estimation Error

To assess estimation error for a given hyperparameter pair, the statistical estimator bias, which is the difference between the estimated PRx value and the “true” value of PRx, was calculated (**eq. 2**). Because each hyperparameter resulted in different time windows, the output of each PRx method was interpolated to compare values directly. For simulated data, where the true value of PRx was known, 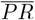 refers to the predefined true PRx value. For patient data, where true PRx is not known, empirical error was calculated by setting the 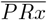 to the average PRx from a range of common hyperparameters (Avg: 5-20 seconds and Corr: 20-50 samples).

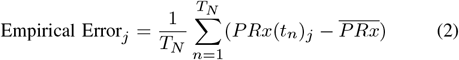

Here, *t*_*n*_ is a time point, *T*_*N*_ is the total time, *j* is the hyperparameter pair, and *PRx*(*t*_*n*_)_*j*_ is the estimated PRx at time *t*_*n*_ for hyperparameter *j*.

#### Standard Deviation of Empirical Error

To assess intrapatient variability impacted PRx estimation error, the standard deviation (SD) of empirical error across all datasets was calculated for a specific hyperparameter *j* (**eq. 3**).

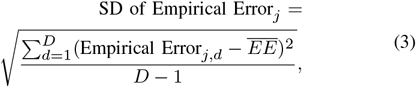

where 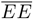 is empirical error (eq. 2) averaged over all data sets (*D*).

#### Noise

To assess the uncertainty or noise for an individual dataset and hyperparameter pair, the standard deviation over time of PRx was calculated.

#### Correlating PRx estimation error and sensitivity with average heart rate

To investigate if average heart rate was associated with PRx estimation error and sensitivity, linear regressions between average heart rate and median PRx, range of PRx values, and empirical error were computed for using all 31 four-hour datasets (from 19 patients).

### Developing Personalized Algorithm for PRx Estimation

To investigate our hypothesis that personalizing the PRx algorithm by averaging over patient heartbeats would provide a more robust and stable estimation of CA, we first identified all heartbeats in ABP signals. Heartbeats were found using a novel sliding peak identification method. Data were segmented into one-minute and one-second windows with one-second overlap. In each window, the beginning of each systolic phase was identified using Matlab’s *findpeaks* function on the inverted ABP segment. To ensure the dicrotic notch was not mistaken as a peak, the minimum peak prominence was set as half of the range of max ABP to min ABP during the minute window (see **Fig. 5a**. for example heartbeat identification). PRx was calculated as before, but the averaging window width was set as a given number of heartbeats rather than seconds.

### Identifying Ideal Hyperparameters

Hyperparameters were identified as ‘ideal’ if they minimized error and SD of error in PRx estimate for both patient and simulated data.

All calculations were conducted in Matlab and are publically available at: https://github.com/jenniferkbriggs/PRxAnalysis.

## III. Results

### PRx is Sensitive to Hyperparameters

Using the development dataset, we calculated PRx for every combination of averaging windows between 1 and 30 seconds and correlation windows between 2 and 65 samples. To validate our PRx calculation, we computed the lagged cross-correlation between the PRx estimate present in the TrackTBI dataset and our PRx estimation using hyperparameters Avg: 10 sec, Corr: 30 samp. (**Supp Fig. 1**). We used a lagged cross-correlation because the PRx calculation is sensitive to the starting time, which is not recorded in the clinical dataset. The average lagged cross-correlation was 0.89, with a standard deviation of 0.05 and an average lag of -0.02 seconds. The primary reasons the cross-correlation was not =1 were this time lag and differences in quality control and artifact removal.

We first compared empirical error using the five common hyperparameters given in **Table I**. The hyperparameter pair Avg: 10 sec, Corr: 40 samp had the lowest average empirical error and smallest SD of empirical error compared to the five common hyperparameters (**Fig. 3a**). Overall, shorter averaging windows (5-6 seconds) were positively biased (hyperparameter overestimates PRx), whereas longer averaging windows were negatively biased (hyperparameter underestimates PRx). These results indicate that hyperparameter choice impacts PRx estimation.

**Fig. 3.**
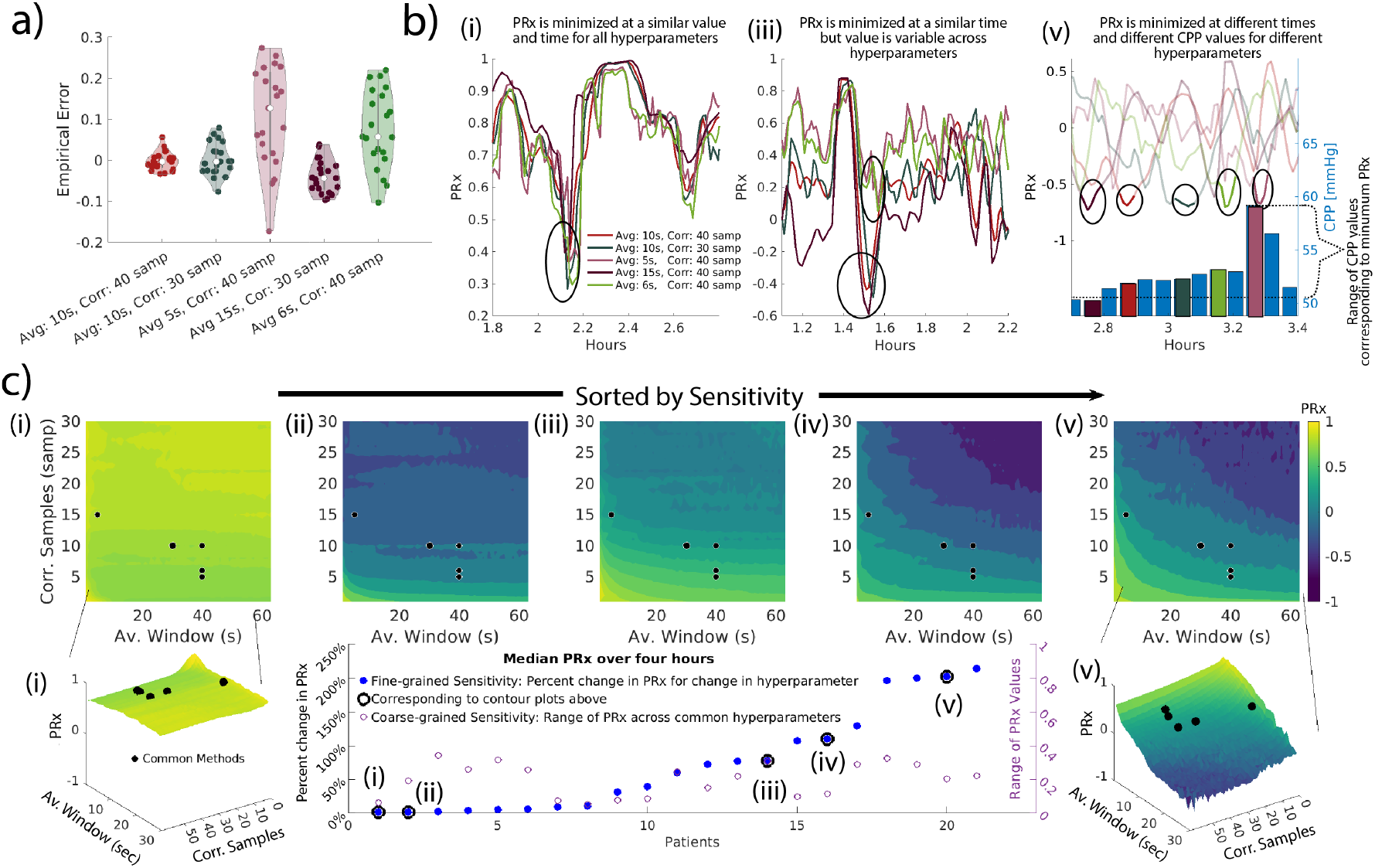
PRx Estimate is Highly Sensitive to Hyperparameters. a) Violin plots showing the PRx estimate empirical error using five common hyperparameters. Data points indicate a four-hour dataset. b) Time courses of PRx for three representative datasets using the five common hyperparameters. The datasets **i, iii, v** correspond to datasets in c. Black circles indicate the minimum PRx estimate. For patient **v**, the PRx estimate is shown on the left y-axis, and CPP is shown on the right axis. CPP bars that correspond to a minimum PRx are colored according to that PRx hyperparameter. The dashed line indicates the range of CPP values where PRx was minimized. c) Example contour plots showing PRx calculation using different hyperparameters. Bottom panel shows 3D contour plots for patient **i** (left) and **v** (right). The scatter plot shows fine-grained sensitivity (left y-axis) and coarse-grained sensitivity (right y-axis) for all datasets, with black circles outlining five datasets corresponding to contour plots.

Clinical decision-making is often based on the CPP value corresponding to minimum PRx[33]. Therefore, we investigated if hyperparameters influence the value of minimum PRx and corresponding CPP. **Fig. 3b** shows one-hour windows when PRx reached a minimum (Roman numerals correspond to datasets in **Fig. 3c**). For dataset **i** (**Fig. 3b**), the value of minimum PRx remained stable and PRx was minimized at similar times (and therefore similar corresponding CPP values) for each hyperparameter. This represents the ideal situation. For dataset **iii**, PRx was minimized at roughly the same time for all hyperparameters, but the estimated PRx value ranged between *functional CA (PRx = -0*.*6) and absent CA (PRx = 0*.*25)*. In dataset **v**, the time of minimum PRx varied by up to 30 minutes, depending on the hyperparameter. Additionally, the CPP (right y-axis in blue bars) corresponding to the minimum PRx ranged from 50-59 mmHg. Therefore, the estimated minimum PRx value and the CPP value corresponding to minimum PRx may be sensitive to hyperparameter choice.

We quantified the fine-grained sensitivity using the average percent change in PRx estimate given a small change in hyperparameter (**Fig 3c** left y-axis of scatter plot). Results ranged from **1.6**% **- 213**% depending on the patient, indicating that a small change in hyperparameters could have a large impact on the PRx estimate. For example, for patient **ii**, using 18 correlation samples and 5, 6, or 7-second averaging windows results in median PRx values of 0.31, 0.28, and 0.17, respectively. These PRx estimates cross the critical threshold of PRx distinguishing ‘intact’ and ‘absent’ CA (=0.25) and will result in different clinical interpretations of the patient’s CA functionality.

We also examined if PRx estimation was variable across the common hyperparameters. Across all analyzed hyperparameters, the PRx estimate ranged between **0.32 and 1.57**. Considering the maximum possible range of PRx values is 2 (PRx can range from -1 and 1), these results correspond to a PRx estimation uncertainty between 16% *-* 79% (**Supp. Fig. 2**). Restricted to the common hyperparameters, the range of the PRx estimate was between **0.05** and **0.34** (**Fig. 3c** right y-axis of scatter plot). This large uncertainty compared to the critical threshold of PRx (=0.25) distinguishing ‘intact’ and ‘impaired’ CA further indicates that the PRx sensitivity to hyperparameters may be large enough to interfere with clinical decision-making.

### PRx estimation is sensitive to intrapatient variability

The large uncertainty in the value of PRx empirical error for each hyperparameter (**Fig. 3a**) indicates that error is influenced by patient variability. Uncertainty in the empirical error was smallest when PRx was estimated using large averaging windows (e.g. empirical error for PRx calculated using Avg: 10 sec, Corr 40 samp had a range of 0.09). Uncertainty in error was largest using small averaging windows (e.g. empirical error for PRx calculated using Avg: 5 sec, Corr: 40 samp had a range of 0.45). We further quantified PRx estimation sensitivity to patient variability. **Fig. 3c** shows contour plots of PRx calculated using all hyperparameter combinations for five representative datasets. The colors, corresponding to different PRx estimates, indicate that PRx is sensitive to hyperparameters, as shown previously. The different morphologies of contours (e.g., horizontal (**ii**) or diagonal and curved (**iv**) and (**v**) indicate that the extent and behavior of PRx sensitivity is heterogeneous across patients. Together, these results indicate that PRx is sensitive to hyperparameters and variability across patients. *Therefore, a patientspecific adjustment to the PRx algorithm may be necessary for ensuring accuracy in PRx estimate for all patients*.

### PRx, PRx Error, and PRx Sensitivity is Associated with Heart Rate

We sought to identify a source of this patient-dependent PRx sensitivity. Averaging in the PRx algorithm was originally meant to remove the influence of heartbeats and respiration[7]. Because heart rate directly influences the number of heartbeats—and consequently, the number of samples—within the averaging window, we examined whether it was associated with sensitivity and error in the PRx estimate. By visual inspection, there is a relationship between average heart rate and error in the simulated dataset (**Fig. 4a**). For patient data, median PRx from all common hyperparameters had a linear relationship with average heart rate (**Fig. 4b**). The range of common hyperparameters was also related to average heart rate (**Fig. 4c**). Finally, while there was a nearly significant linear relationship (p=0.08) between average heart rate and empirical error for hyperparameter pair Avg: 5 sec, Corr: 40 samp, this relationship was not present for other hyperparameter pairs (e.g. Avg: 10 sec, Corr: 40 samp) (**Fig. 4d,e**). We did not identify any linear relationships between heart rate variability and PRx errors. These results imply that heart rate is related to PRx sensitivity, indicating that there may be a way to reduce error by reparameterizing the PRx algorithm to heartbeats.

**Fig. 4.**
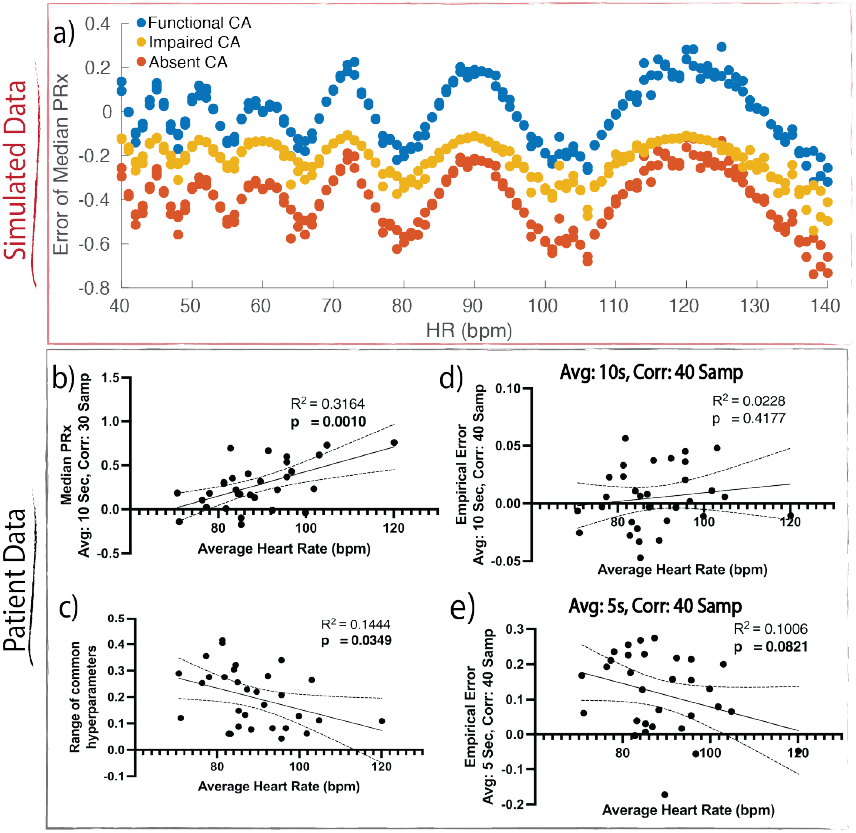
PRx Depends on Heart Rate. a) Scatter plot showing relationship between error in median PRx and heart rate for three levels of CA function in simulated data. b-e) Scatter plots relating average heart rate to different PRx evaluation metrics. The black line indicates linear regression with 95**%** confidence intervals shown as dashed lines. Bolded p-values indicate statistical significance or near significance.

### Personalized PRx Algorithm (pPRx) Results in Reduced Error, Sensitivity to Hyperparameters, and Noise

We corrected for heart rate by identifying all heartbeats in the patient and simulated data and averaging over heartbeats rather than seconds (**Fig. 5a**). For example, if the original hyperparameter pair was Avg: 5 sec, Corr: 40 samp, we instead averaged over five heartbeats and then correlated over 40 of these averaged samples (**Fig. 5a**). We call this new heartbeat-based algorithm personalized PRx algorithm (**pPRx**) and the standard seconds-based algorithm **sPRx**. The pPRx calculation did not notably impact computation time, which is important for real-time computation of PRx in operational settings. We compared the performance of sPRx and pPRx using simulated and patient data in the development and validation datasets. For simulated data, pPRx was less sensitive to hyperparameters and intrapatient variability and had a lower average estimation error than sPRx (**Fig. 5b, Table V**). For patient data, pPRx decreased the noise of PRx estimation for both datasets (**Table V, Fig. 5c,d**,). The pPRx calculation was less sensitive to hyperparameters than pPRx (**Fig. 5e, Table V**) and did not have a notable impact on average empirical error (**Fig. 5f, Table V**). The pPRx calculation decreased the standard deviation of empirical error, which corresponds to patient dependent errors for both datasets (**Table V**).

**Fig. 5.**
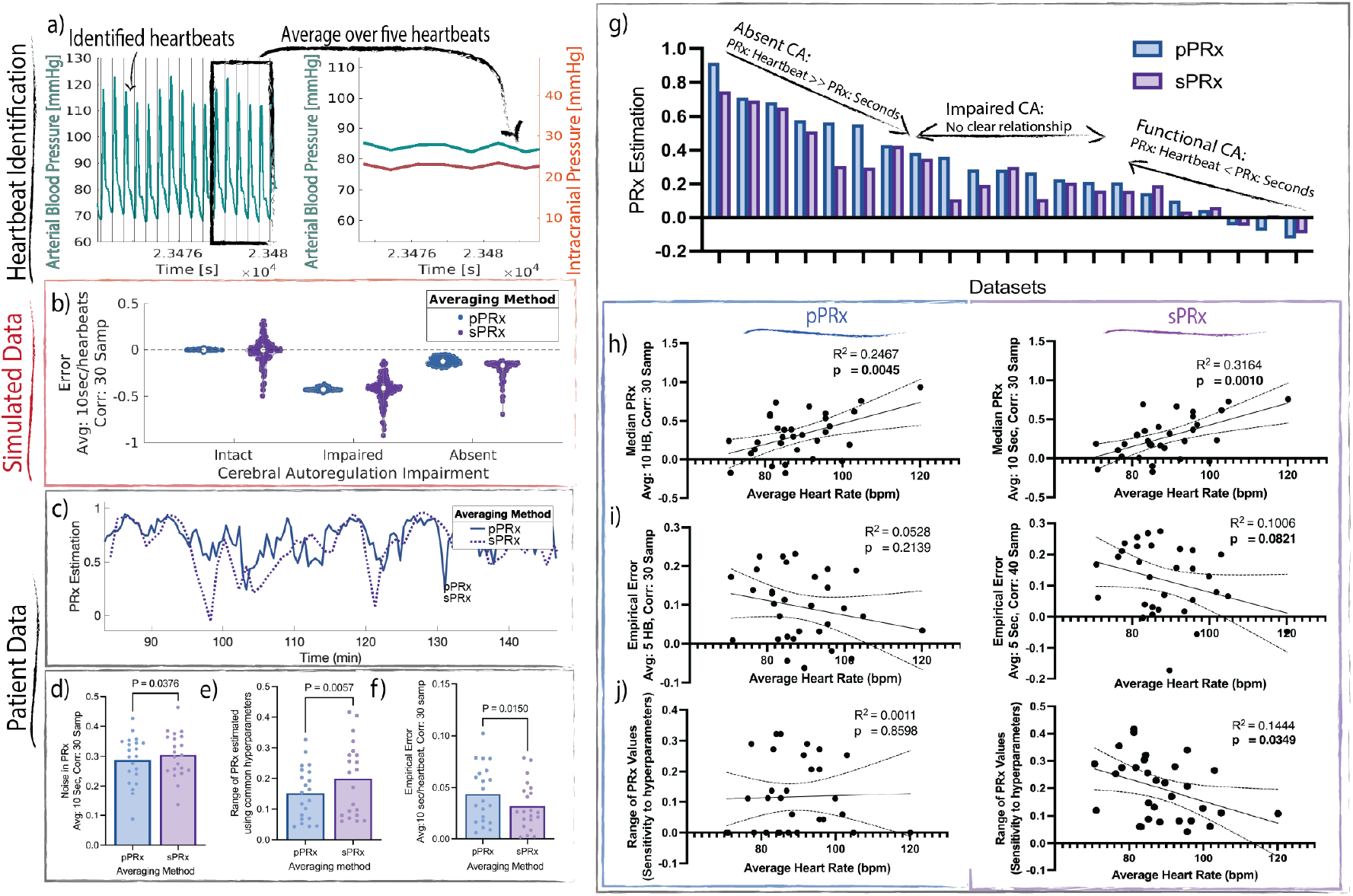
Personalized PRx (pPRx) is More Robust to Hyperparameters and Patient Variability than sPRx. a) Methodological schematic showing representative ABP signal from patient data with black lines indicating heartbeat start and end times (left). ABP (teal) and ICP (red) are averaged over a given number of these heartbeats (right). b) Violin plots comparing estimation error in pPRx (blue) and sPRx (purple) in simulated data. c) Representative time series from patient data of pPRx (blue) and sPRx (purple). d-f) comparisons of the performance of the pPRx (blue) and sPRx (purple) for 21 development datasets using one-tailed paired t-tests. Dots indicate a single dataset. g) Bar plot showing PRx value for all patient data sets. h-j) Correlations between average heart rate and PRx metrics calculated for using pPRx (left) and sPRx (right). Solid black line shows linear fit with dashed lines showing 95**%** confidence intervals.

**TABLE V.** The Personalized PRx Algorithm Decreases Sensitivity to Hyperparameters and Patient Variability, Error in PRx, and Noise Compared to Standard Algorithm. Bold value* indicates which method had better performance compared within datasets. Numbers represent the average metric value across all datasets and the five common hyperparameters. SD indicates standard deviation. Max Empirical Error is the maximum *absolute value* of empirical error

|  | sPRx |  | pPRx |  |
| --- | --- | --- | --- | --- |
|  | Development | Validation | Development | Validation |
| *Sensitivity of PRx to Hyperparameters* |  |  |  |  |
| Average Range of PRx Error (simulated) | 0.17 | 0.11 | **0.06\*** | **0.08\*** |
| — Intact CA | 0.15 | 0.07 | **0.00\*** | **0.00\*** |
| — Impaired CA | 0.18 | **0.11\*** | **0.06\*** | 0.13 |
| — Absent CA | 0.19 | 0.16 | **0.11\*** | **0.00** |
| Coarse-grained Sensitivity (patient) | 0.20 | 0.19 | **0.16\*** | **0.18\*** |
| Fine-grained Sensitivity (patient): | 74.09% | 44.73% | **55.21%\*** | **39.66%\*** |
| *Sensitivity of PRx to Patient Variability* |  |  |  |  |
| SD of Error (simulated) | 0.24 | 0.19 | **0.18\*** | **0.16\*** |
| SD of Empirical Error (patient) | 0.06 | 0.05 | **0.05\*** | **0.04\*** |
| *PRx Estimation Error* |  |  |  |  |
| Average Error (simulated) | -0.23 | -0.22 | **-0.18\*** | **-0.19\*** |
| — Intact CA | -0.01 | -0.02 | **0.00\*** | **0.00\*** |
| — Impaired CA | -0.43 | -0.43 | **-0.41\*** | **-0.38\*** |
| — Absent CA | -0.25 | -0.38 | **-0.13\*** | **-0.17\*** |
| Max Empirical Error (patient) | 0.27 | 0.27 | **0.23\*** | **0.23\*** |
| Average Empirical Error (patient) | **0.03\*** | 0.05 | **0.03\*** | **0.04\*** |
| Noise (patient) | 0.30 | 0.31 | **0.29\*** | **0.30\*** |

The average value of PRx was not noticeably different between the two averaging methodologies (**Fig. 5g**), indicating that the interpretation of the sPRx likely extends to the pPRx. For simulated data, sPRx had the smallest error for intact CA phenotypes and underestimated the true PRx value for absent and impaired CA phenotypes (**Fig. 4a, Table. V**). The pPRx calculation decreased this CA functionality-specific error in simulated data (**Fig. 4b**). A similar trend was present using patient data - for patients absent CA (PRx *>*0.25), pPRx was larger on average than sPRx (**Fig. 5g**.).

We evaluated if pPRx affected the previously observed relationship between PRx and heart rate. The pPRx calculation did remove the relationship between empirical error (using hyperparameters Avg: 5 sec, 40 samp) and average heart rate for PRx (**Fig. 5i**.) and between the range of common hyperparameters and average heart rate (**Fig. 5j**.), but not between median PRx and average heart rate (**Fig. 5h**). Together, these results indicate that the pPRx calculation removes heart rate-associated PRx sensitivity and error without greatly altering the PRx estimate.

### Ideal Hyperparameters in Standard Methodology Improve PRx Estimate

We also investigated which hyperparameters in the standard algorithm decreased PRx errors. Hyperparameter pairs in averaging range: 9-10 seconds and corelation range: 35-55 samples minimized the standard deviation (SD) of error and average error in simulated and patient data (**Fig 6a-d**) in the development dataset. Hyperparameter pair: Avg: 10 seconds and Corr: 40 samp, which we refer to as *the ideal hyperparameter pair*, was the only common hyperparameter pair (**Table I**) that fit within this range. This ideal hyperparameter pair significantly decreased the standard deviation of empirical error compared to other common hyperparameters in both the development and validation datasets (**Fig 6e-h**). Notably, the ideal hyperparameter pair outperformed the most common hyperparameter pair (Avg: 10 sec, Corr: 30 samp) in most metrics (**Supp. Table I, Fig 6e-h**).

**Fig. 6.**
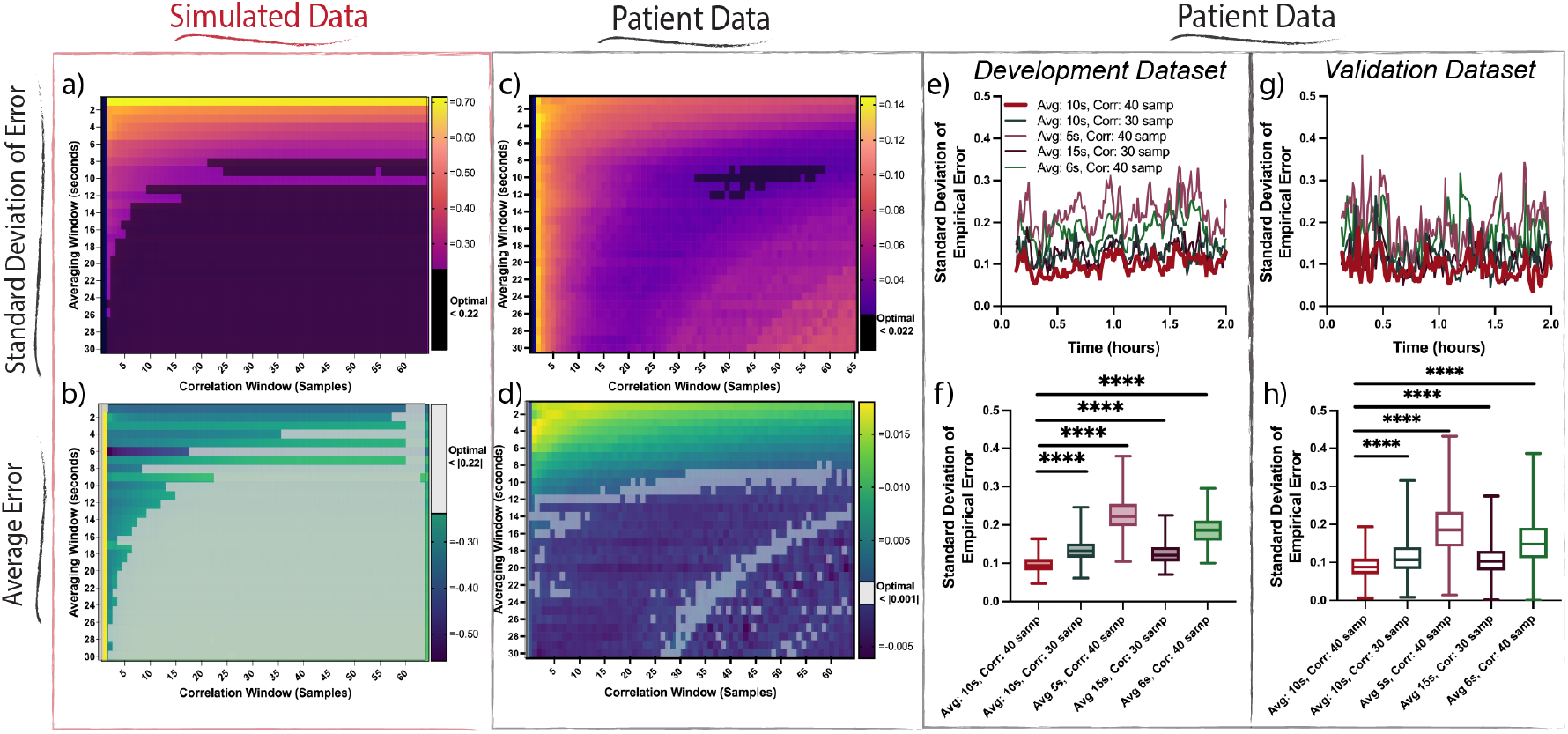
10 Second Averaging Window and 40 Sample Correlation Window Improves sPRx Estimate. a) Contour plots showing standard deviation (SD) of error in sPRx for simulated data for many hyperparameter pairs. Black shading indicates hyperparameter pairs resulted in a minimum SD of error. b) Contour plots showing average error in simulated data. White indicates hyperparameter pairs resulted in a minimum error. c) As in a but for patient data. d) As in c, but for patient data. e) One-hour time series of the standard deviation of empirical error across all patients in the development data set for the five common hyperparameters. The hyperparameter pair that minimizes the standard deviation of empirical error is bolded (red). f) Comparison between the standard deviation of empirical error across all patients in the development data set for each time point. The averaging window of 10 seconds and correlation window of 40 samples had a significantly smaller error (p***<***0.0001) than other common hyperparameters. g) As in e for the validation dataset. h) As in f for the validation dataset. Analysis was paired one-way ANOVA with multiple comparisons.

## IV. Discussion

Neurological injuries are a leading cause of death throughout the world. The pressure reactivity index (PRx) is a metric used to evaluate cerebral autoregulation (CA) function and guide clinical decision-making for patients in neurocritical care[5], [7], [8]. PRx has high variance (e.g. noise) in time[34] and across clinical monitors[35], and can be confounded by extra-physiologic variables, such as hospital bed type[36]. The objectives of this study were to enhance the robustness of PRx by personalizing the algorithm to remove errors and by identifying ideal hyperparameters in the standard algorithm.

### Recommendations Based on Findings

*When possible, we recommend using the pPRx algorithm to quantify CA. The pPRx algorithm showed decreased noise and sensitivity to intrapatient variability for two independent datasets, allowing for greater reliability of PRx and its use for clinical decision support (****Fig. 5, Table V***). The code for this new algorithm is publicly available (see methods).

In the absence of pPRx, we recommend using averaging windows between 9-10 seconds and correlation windows between 40-55 samples. These ideal hyperparameters decrease estimation error and sensitivity to intrapatient variability in the development and validation datasets (**Fig. 6, Supp. Table I**) compared to the most common hyperparameter pair (Avg: 10 sec, Corr: 30 samp).

### Hyperparameter Sensitivity

A central challenge in studying PRx is that it cannot be directly validated due to the invasive nature of ICP measurement and the lack of a ‘gold standard’ global continuous CA evaluation metric. Previous studies have shown that PRx is a more successful predictor of mortality than other CA metrics[37] and is a good predictor of the lower limit of autoregulation in animals[38], [39]. However, quantifying true PRx error is not possible with patient data. Therefore, we took two approaches to evaluate the PRx estimation. First, we used patient data, and quantified PRx sensitivity using multiple empirical metrics (**Table IV**). Second, we created simulated data to quantify PRx estimation error directly. Using these approaches, we identified that the PRx algorithm is sensitive to to hyperparameters and intrapatient variability.

The sPRx algorithm was sensitive to hyperparameters may impact clinical interpretation of CA function and clinical decisions. On average, changing hyperparameters caused the sPRx estimate to span nearly 50% of all possible values (**Supp. Fig. 2**). Additionally, the average sensitivity to hyperparameters (0.2) was nearly as large as the critical PRx threshold distinguishing between ‘intact’ CA and ‘absent’ CA (0.25)[19], [40]. A primary way that PRx is used to guide clinical decision-making is through targeting the CPP that corresponded to minimized PRx occurred [21], [33]. In our study, this corresponding CPP was also sensitive to hyperparameters (**Fig. 3b**). However, this minimum PRx is often identified using an average parabolic fit, which may increase robustness.

Our results also indicate that sPRx error is dependent on variability across patients. The range of empirical error, which indicates how much patient variability influences error, was near to, or larger than the critical threshold for all five common hyperparameter choices (**Fig 3a**), indicating that patient variability may influence sPRx enough to interfere with its clinical interpretability.

### Personalizing the Pressure Reactivity Index

We identified heart rate as a major variable related to standard PRx (sPRx) estimation sensitivity (**Figs 3, 4**). This sensitivity has direct clinical implications. Heart rate variability and average resting heart rate are associated with age, ethnicity, and sex even after controlling for other health factors[41], [42]. Therefore, sPRx may contain different errors for certain patient demographics, particularly pediatrics who have higher resting heart rates than the adult population.

We developed the personalized pressure reactivity index (pPRx) by reparameterizing the averaging window to heartbeats rather than seconds to remove this heart rate-dependent bias. On average, pPRx was less sensitive to hyperparameters and intrapatient variability and had lower noise compared to sPRx (**Fig. 4, Table V**).

We believe the improved accuracy of pPRx results from natural variations in heart rate impacting the robustness of sPRx. First, in the sPRx calculation, changes in heart rate can alter the number of heartbeats within the averaging window, introducing variability in the computed average. Second, when parameterizing by time, the averaging window likely a non-integer number of heartbeats, resulting in incomplete pressure waveforms and introducing bias in the computed average. The pPRx calculation ensures that each window contains an equal number of complete pressure waveforms, independent of changes in heart rate.

By ensuring an equal number of heartbeats, pPRx is more robust to changes in heart rate, which can rapidly change in critical care settings. Because sPRx error depends on heart rate, a sudden change in patient state could increase estimation error. Alternatively, since pPRx identifies heart rates in real-time, a change in the patient’s state will be immediately accounted for, making pPRx potentially more reliable in critical care settings. There are likely similar parameterizations that could enhance the robustness of other CA-based metrics, such as CPP-opt, Mx, or ORx[21], [43], [44].

As opposed to sPRx, pPRx did not show a statistically significant linear relationship between average heart rate and PRx errors (**Fig. 5 h,i**). This is expected because the pPRx accounts for the computational influence of heart rate on the computed average. However, average heart rate and median PRx was still linearly correlated using pPRx, possibly indicating a physiological relationship between PRx and average heart rate. CA function and PRx (calculated using sPRx method) have been associated with heart rate-related factors, such as age, disease state, and vascular comorbidities (see review articles [5], [45]). Additionally, because heart rate directly influences cardiac output and ABP, it is possible that heart rate directly influences CA function or that impaired CA triggers compensatory mechanisms, such as heart rate, to increase cerebral perfusion. Further investigation into the physiological relationship between average heart rate and CA functionality is warranted.

### Identifying the Ideal Hyperparameter Pair

One response to the finding that sPRx calculation is highly sensitive to hyperparameters is to agree on a single, ideal hyperparameter pair to use across all applications. Hyperparameter pair of Avg: 10 sec, Corr: 40 samp was the most robust to intrapatient variability (**Fig. 6f,g**). Correspondingly, there was not a linear relationship between heart rate and PRx empirical error calculated using this ideal hyperparameter pair, whereas there was a nearly significant relationship for other hyperparameter pairs (e.g. Avg: 5 sec, Corr: 40 samp) (**Fig. 4d,e**). This finding supports our hypothesis that variability in heart rate is a factor underlying the sensitivity of sPRx error to intrapatient variability.

However, defining an ideal hyperparameter pair does not remove sPRx’s sensitivity to hyperparameters, which also indicates sensitivity to missing data. For example, assume the averaging window for sPRx is ten seconds, but there are two seconds of data missing. This ten second averaging window will only contain eight seconds worth of data and will result in the same computed average as an eight second averaging window. Therefore, in the sPRx algorithm, missing data has a similar effect as changing hyperparameters.

Missing data are common in clinical settings, making sensitivity to missing data will be clinically impactful. For example, in **Fig. 3c patient ii**, changing the averaging window by one second, which corresponds to missing one second of data per averaging window changes the median PRx estimation from 0.17 to 0.28, which crosses the critical threshold and changes the clinical interpretation of CA functionality. More broadly, in real clinical settings missing data are common, highly complex, and are often a function of the external environment via the healthcare process [46], [47]. The examples above do not fully characterize pPRx and sPRx robustness to data missingness. Such an analysis is an important topic for future work.

In this study, we did not include time series with large amounts of missing data, and our sample size was relatively small. Therefore, our results are likely a conservative estimate of the possible error and sensitivity of PRx (because the prevalence of outliers generally increases as sample size increases). Further analysis should be conducted on larger cohorts.

## V. Conclusion

The pressure reactivity index (PRx) is an important metric for assessing cerebral autoregulatory function and aiding clinical decisionmaking for neurocritical care patients. Here, we show that the PRx calculation is sensitive to hyperparameters and intrapatient variability, partly due to variability in average heart rate. Reducing this sensitivity is crucial for increasing the usefulness of PRx, particularly when PRx is applied to demographic groups with different average heart rates and heart rate variability. Therefore, we developed a new personalized heartbeat-specific PRx algorithm (pPRx) that *reduces error, noise, patient sensitivity, and hyperparameter sensitivity compared to the standard method*.

## Data Availability

All simulated data and code will be made available at: https://github.com/jenniferkbriggs/PRxAnalysis.git
Patient data can be requested by contacting TRACK-TBI.

https://tracktbi.ucsf.edu/transforming-research-and-clinical-knowledge-tbi

## VI. Supplement

**Fig. S1.**
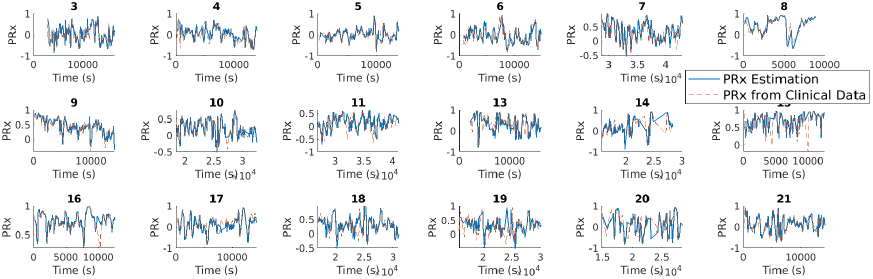
Validation of PRx against clinical PRx datasets. Blue indicates our lagged PRx estimate. Red represents PRx estimate output by clinical PRx datasets.

**Fig. S2.**
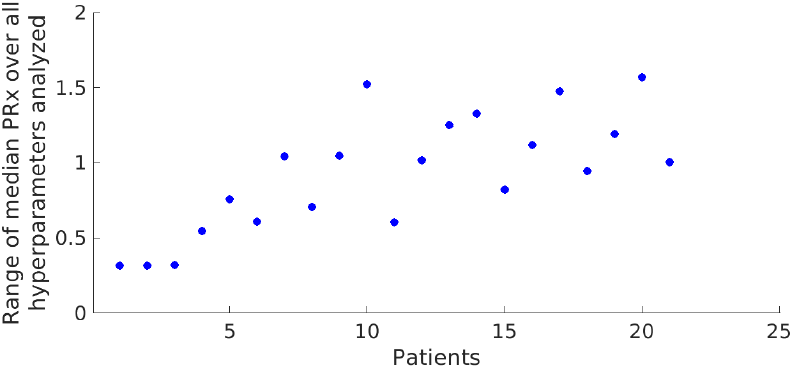
Range of median PRx values over all hyperparameters analyzed.

**TABLE S1.**
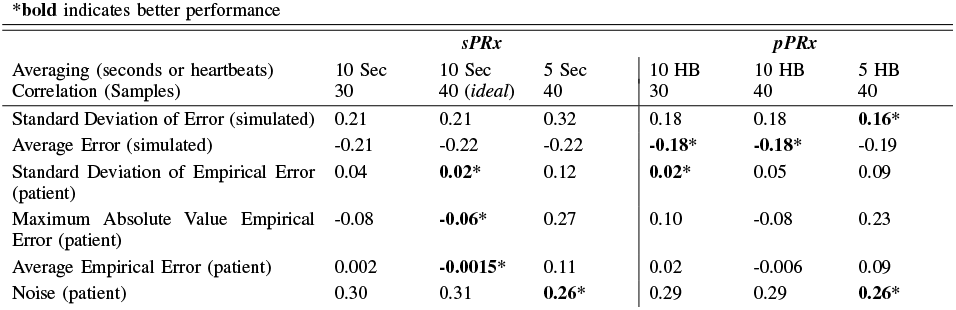
Comparing the Performance of the Personalized Pressure Reactivity Index (pPRx) and Standard Pressure Reactivity Index (sPRx) for Three Different Hyperparameters. Bold value* indicates the method and hyperparameter pair that had the best performance. Metrics are assessed for the development dataset only. Numbers represent the average metric value for all datasets.

## Notes

### Competing Interest Statement

The authors have declared no competing interest.

### Funding Statement

This study was funded by National Institutes of Health R01s LM006910 and LM012734 to DJA, and National Science Foundation (NSF) Graduate Research Fellowship DGE-1938058 Briggs to JKB

### Author Declarations

The ethics committee/IRB of the respective institutions enrolled in TRACK-TBI gave ethical approval for this work.

### Summary of Updates

INclude statement of possible publication in IEEE journal

